# Supervised Contrastive Learning-based Digital Biomarker Discovery for Wearable IMU Gait Signals

**DOI:** 10.64898/2026.07.02.26357115

**Authors:** Seyed Mehdi Mohtavipour

## Abstract

Wearable inertial measurement units (IMUs) provide a practical and objective approach for gait assessment in clinical populations. Although several handcrafted gait features have been proposed, these features may not fully capture the multidimensional signal characteristics associated with different pathological gait patterns. This study proposes a digital biomarker called Embedding-Distance Gait Biomarker (EDGB) based on supervised contrastive representation learning of wearable IMU signals. A compact multi-input convolutional neural network is developed to encode raw acceleration, angular velocity, and their temporal derivatives into a 32-dimensional latent representation. Class-specific prototypes are computed from the training embeddings of healthy, neurological, and orthopedic participants. The proposed EDGB is then derived from the distances between each trial embedding and the learned group prototypes. The proposed architecture is evaluated on the publicly available Voisard clinical gait dataset using a subject-level split, with 20% of participants held out for testing to prevent leakage across repeated trials. On unseen test subjects, the proposed biomarker distinguished healthy from neurological, healthy from orthopedic, and neurological from orthopedic gait patterns with AUCs of 90.59%, 88.47%, and 99.50%, respectively. The biomarker also demonstrated a large group effect, with clinical category explaining 71% of its variance. Reliability analysis showed significant consistency across repeated trials, with an ICC (2,1) of 0.82, indicating that most variability reflected between-subject differences rather than within-subject trial-to-trial fluctuations.

## 1. Introduction

Gait is a fundamental component of human mobility and serves as an important indicator of neurological, musculoskeletal, and general health status [1]. Alterations in gait are common across a wide range of neurological and orthopedic conditions, including Parkinson’s disease (PD) [2], cerebrovascular accident (CVA) [3], peripheral neuropathy [4], osteoarthritis [5], and anterior cruciate ligament (ACL) injury [6]. These disorders often affect different aspects of locomotion, such as walking speed, balance, coordination, rhythm, symmetry, and movement smoothness. Consequently, quantitative gait assessment has become increasingly important for disease diagnosis, severity evaluation, treatment monitoring, and longitudinal follow-up [7]. Traditionally, gait assessment in clinical practice relies on visual observation by trained clinicians or on specialized motion-capture laboratories. While expert clinical assessment remains valuable, visual evaluation is inherently subjective and may suffer from inter-rater variability. In contrast, laboratory-based motion capture systems provide highly accurate measurements but are expensive, time-consuming, and difficult to deploy outside specialized facilities. These limitations have motivated the development of wearable sensing technologies, particularly inertial measurement units (IMUs), which enable objective, portable, and cost-effective gait analysis in both clinical and real-world settings [8].

Recent advances in wearable sensing have led to the collection of large datasets containing synchronized accelerometer and gyroscope recordings from healthy individuals and patients with neurological and orthopedic disorders. Such datasets provide an opportunity to investigate disease-specific gait signatures and develop digital biomarkers capable of objectively characterizing locomotor impairments. Conventional gait analysis has traditionally relied on handcrafted features derived from biomechanical and clinical knowledge. These features provide direct interpretability and can often be associated with specific motor impairments. Previous studies have described gait using clinically meaningful domains such as pace, rhythm, variability, asymmetry, and postural stability [9]. Accordingly, spatiotemporal measures such as stride time, step length, and gait cadence have been widely used to quantify gait alterations across different patient populations [10]. However, although these measures are clinically interpretable, each feature typically captures only a limited aspect of gait and may not fully represent the multidimensional nature of gait impairment. Motivated by these limitations, machine learning and deep learning approaches have increasingly been applied to gait classification and disease identification [11,12]. Deep neural networks can learn complex spatiotemporal patterns directly from raw sensor signals and have shown promising performance in several gait-related tasks. Despite their predictive capability, these models often provide limited interpretability and may function as black-box classifiers. In addition, reported performance can be strongly affected by the evaluation protocol [13]. In particular, trial-wise train-test splits may introduce information leakage when repeated trials from the same participant appear in both training and testing sets, leading to overly optimistic estimates of generalization. Therefore, subject-wise evaluation is essential for clinical applications to ensure that models learn disease-related gait characteristics rather than subject-specific movement signatures.

The availability of clinically annotated multi-pathology gait datasets creates a unique opportunity to systematically investigate gait characteristics across different movement disorder populations. In this study, we use the Voisard dataset [14], which provides wearable IMU recordings from healthy participants and patients with neurological and orthopedic conditions. The dataset includes gait signals collected from multiple body-worn sensors during a standardized walking protocol, together with clinical labels and metadata. This structure makes it suitable for developing and evaluating data-driven gait representations that can distinguish between healthy, neurological, and orthopedic gait patterns while preserving clinical relevance. This study proposes an Embedding-Distance Gait Biomarker (EDGB) derived from supervised contrastive learning of multi-sensor IMU signals. As illustrated in Fig. 1, acceleration and angular velocity signals acquired from head, lower back, right foot, and left foot IMUs are first preprocessed and augmented, and their corresponding derivative signals are computed to capture higher-order movement dynamics. These signals are then provided to a compact CNN-based encoder network that learns a low-dimensional gait representation. The proposed EDGB is subsequently constructed using the distances between each gait embedding and learned prototype vectors. Through this framework, the present study aims to investigate whether a learned gait representation can provide a discriminative digital biomarker for healthy, neurological, and orthopedic gait patterns.

**Figure 1.**
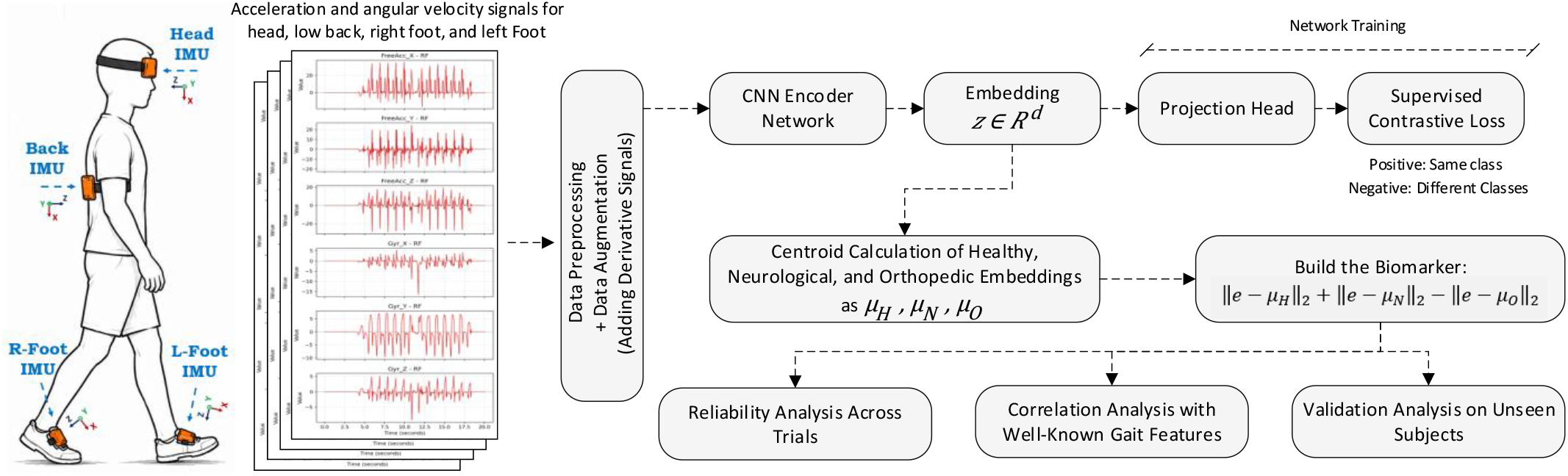
Overall architecture of proposed digital biomarker discovery based on the wearable IMU gait signal

## 2. Related Works

Several previous studies have investigated feature-based representations for gait analysis. Authors in [15] used IMU-based gait data for frailty assessment in older adults and evaluated handcrafted features such as walking distance, total walking time, number of strides, stride length, and stride time for training on a random forest machine leaning algorithm. The test accuracy of using the handcrafted features in study is reported 63.3%. A quantitative gait assessment framework is proposed in [16] using two IMU sensors on right and left foots, extracted 17 spatial and temporal features. It was demonstrated that with only these two sensors, their index feature can distinguish between healthy subjects and subjects with lower-limb injury history. Another gait pattern identification is introduced in [17] using explicit gait feature points extracted from superimposed gait cycles. Their work extracted stance time, swing time, gait time, cadence, and three characteristic gait points corresponding to heel strike, mid-stance, and toe-off, showing that a small number of interpretable gait features can distinguish individual walking patterns. To differentiate early-onset ataxia, developmental coordination disorder, and healthy controls, authors in [18] introduced gait feature fusion network suggesting a potential tool for objective assessment compared to semi-quantitative clinical rating scales. Another IMU-based gait recognition using five body-worn inertial sensors and multi-sensor fusion is introduced in [19]. Their study highlighted the importance of sensor location and fusion strategy, showing that combining information from multiple IMUs can improve gait-identification performance compared with single-sensor analysis. These studies demonstrated handcrafted gait features provide interpretable and clinically meaningful descriptors; however, they showed important limitations in capturing full aspects of complex multidimensional gait abnormalities.

To overcome the limitations of conventional gait measures, recent studies have increasingly adopted deep learning approaches that learn discriminative representations directly from raw IMU signals. These methods eliminate the need for manual feature engineering and can automatically capture complex temporal and spatial relationships within gait data. For example, authors in [20] proposed a deep learning framework for gait event detection using a single shank-mounted IMU and demonstrated accurate identification of heel-strike and toe-off events in both healthy individuals and neurological patients. Their results highlighted the capability of deep neural networks to learn gait events directly from inertial signals with a precision more than 97%. Another work in [21] introduced a deep learning approach for detecting freezing-of-gait episodes in Parkinson’s disease patients using wearable IMUs. By employing convolutional neural networks on raw sensor data, the method achieved robust detection of freezing events and demonstrated the potential of deep learning for monitoring disease-specific gait abnormalities. A convolutional recurrent neural network (CRNN) that combined CNN-based feature extraction with recurrent temporal modelling is introduced in [22] to classify locomotion modes, gait phases, and transitions from IMU recordings. Their work demonstrated that hierarchical deep architectures can effectively capture both local motion patterns and long-term temporal dependencies. Another usage of deep learning in IMU signal is presented in [23] for fall-risk assessment in older adults, where neural networks were developed to distinguish fallers from non-fallers. Some studies have also investigated deep learning for extracting gait characteristics and subject-specific walking patterns. For example, Deep learning models in [24] have been applied to estimate gait parameters from leg- and shoe-mounted IMUs, demonstrating the ability of neural networks to learn relationships between raw inertial signals and clinically relevant gait measures. There are also some other works that transformed IMU signals into time-frequency representations such as spectrograms and employed convolutional neural networks for gait recognition and gait classification [25]. Although these deep learning approaches have demonstrated strong predictive performance, most of them are designed for event detection or task-specific classification. In contrast, the present study aims to develop a digital gait biomarker by embedding complex, multidimensional IMU gait signals into a latent representation space. This representation is designed not only to distinguish gait patterns across movement disorder categories, but also to provide a compact biomarker that can be used to clinically assess and quantify the performance of subjects.

## 3. Materials and Methods

### 3.1 Dataset, Participants, and Experiment details

This work employed the publicly available Voisard gait dataset [14], which contains wearable inertial measurement unit (IMU) recordings collected from healthy individuals and patients with neurological and orthopedic disorders. The dataset comprises 1,356 gait trials acquired from 260 participants during a standardized walking protocol consisting of standing still, walking 6–10 m, performing a 180° U-turn, walking back to the starting position, and stopping. The study population consisted of three major groups: healthy controls (73 participants, 360 trials), neurological patients (143 participants, 784 trials), and orthopedic patients (44 participants, 212 trials). As shown in figure 1, participants were equipped with four synchronized IMUs positioned on the forehead, lower back (L4/L5), and the dorsal aspect of both feet. Sensor data were recorded at 100 Hz and included three-axis acceleration and three-axis angular velocity signals. For each trial and for each IMU sensor, three-axis acceleration and three-axis angular velocity are provided. The dataset additionally includes demographic information, gait-event annotations, and U-turn boundaries, enabling for direct conventional gait measures.

### 3.2 Deep Learning Architecture

This section describes the proposed deep learning architecture for learning gait representations from wearable IMU signals. The inputs to the network consist of the magnitude of acceleration denoted by ‖*α*(.)‖, magnitude of angular velocity denoted by ‖x*ω*(.)‖, derivative of magnitude of acceleration denoted by ‖*j*(.)‖, and derivative of magnitude of angular velocity denoted by ‖*α*(.)‖, acquired from the head (HE), lower back (LB), left foot (LF), and right foot (RF) sensors. In addition, the first temporal derivatives of these signals, corresponding to jerk and angular acceleration, are included as supplementary inputs. Consequently, the network receives a total of 16 input signals, comprising acceleration magnitude, angular velocity magnitude, jerk magnitude, and angular acceleration magnitude from each of the four IMUs. The inclusion of derivative signals provides explicit information about temporal changes in movement dynamics. By supplying these higher-order motion descriptors directly to the network, informative features can be extracted using a compact architecture with fewer trainable parameters compared with relying solely on raw signals. The magnitude of acceleration and angular velocity is computed from the three orthogonal sensor axes as:

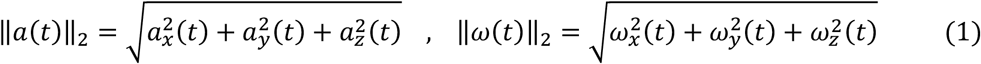

Where *x, y,z* represent three sensor axes. Since the duration of gait recordings varies across participants, all input signals are resampled to a fixed length of 1900 samples prior to network training. Given the acquisition frequency of 100 Hz, this corresponds to approximately 19 seconds of gait data. The proposed architecture is illustrated in Fig. 2. The feature extraction stage consists of three one-dimensional convolutional layers with 4, 8, and 16 output channels, respectively. The first convolutional layer applies four filters with a kernel size of 5 and padding of 2 to the 16-channel input signal. The second convolutional layer employs the same kernel size and padding, while the third convolutional layer uses a kernel size of 3 and padding of 1. Batch normalization is applied after each convolutional layer to improve training stability and convergence by learning channel-wise scaling and shifting parameters. Rectified Linear Unit (ReLU) activations are used throughout the network. Furthermore, max-pooling with a pooling factor of 2 is applied after the first and second convolutional layers to progressively reduce the temporal resolution while preserving salient gait characteristics. To capture both global and localized temporal information, the output feature maps of the final convolutional layer are processed using adaptive average pooling and adaptive max pooling. Both operations generate 16 feature maps distributed across 8 temporal bins. The resulting pooled representations are concatenated and flattened, producing a compact representation that preserves complementary statistical information from the gait sequence. This representation is described as follows:

**Figure 2.**
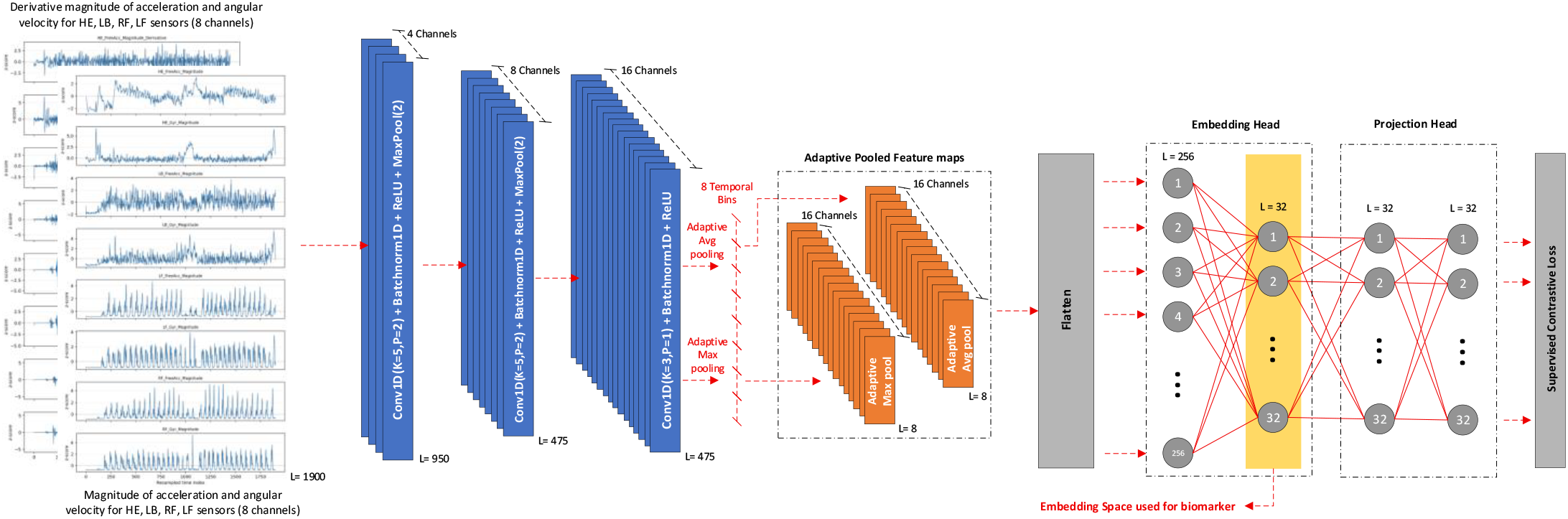
Proposed network architecture based on the supervised contrastive learning, inputs are 8 IMU signals of acceleration and angular velocity magnitude for HE, LB, RF, LF sensor locations and 8 corresponding derivative signals, final embedding vector in the embedding layer with the length of 32 is selected as biomarker

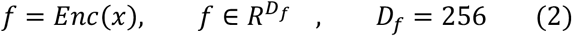

Where *f* denotes the flattened feature vector obtained after adaptive average and adaptive max pooling and *x* is the input channels. Following the [26] framework, the fully connected stage consists of an embedding network and a projection network. The embedding network transforms the extracted convolutional features into a 32-dimensional latent representation, which serves as the proposed digital biomarker. Subsequently, the projection network maps the embedding into a contrastive feature space used exclusively during supervised contrastive learning. The embedding and projection latent representation are given as follows:

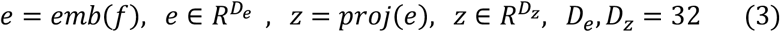

Both *e* and *z* are 32-dimentional vectors of latent representations. During inference, only the 32-dimensional embedding vector is retained and used for biomarker computation and gait pattern analysis.

### 3.3 Supervised Contrastive Loss

Training of the architecture shown in Fig. 2. aims to learn an embedding space in such a way that gait recordings belonging to the same clinical category are clustered together, while recordings from different clinical categories are separated. To achieve this objective, the Supervised Contrastive Learning (SupCL) framework proposed in [27] is employed. Unlike conventional classification losses, SupCL explicitly utilizes class labels to construct positive and negative sample pairs within each batch. The learned representation is therefore encouraged to maximize similarity between samples of the same class while increasing the distance between samples belonging to different classes.

The Voisard dataset contains three clinical categories, namely healthy, neurological, and orthopedic participants, with multiple gait trials available for each subject. Consequently, positive samples can be naturally constructed from gait trials belonging to the same clinical category, whereas samples from different clinical categories are treated as negatives. During training, the encoder and projection network are jointly optimized such that embeddings from the same category form compact clusters in the latent space, while embeddings from different categories become increasingly separated. The SupCL loss is defined as follows:

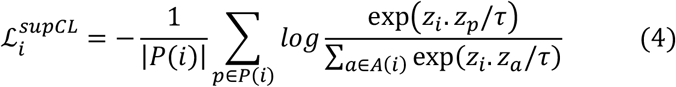

Where *P(i)* represents all positive samples in the batch that have the same clinical category as sample *i*, excluding sample *i* itself. *A(i)* represents all other samples in the batch, excluding sample *i*, and therefore includes both positive samples from the same category and negative samples from different categories. *z*_*i*_, *z*_*p*_, *z*_*α*_ are the latent projection vectors of the anchor sample, positive samples, and all comparison samples, respectively. τ is the temperature parameter, which was set to 0.1 in this study. This loss pulls embeddings of samples from the same clinical category closer together and pushes embeddings of samples from different clinical categories farther apart. The SupCL loss for one batch is obtained by averaging the anchor-level losses over all samples:

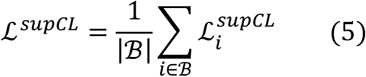

Where *B* is the contrastive batch, and | *B* | is the number of samples in the batch.

### 3.4 Embedding Distance Gait Biomarker (EDGB)

After training the SupCL framework, the projection head is discarded and only the 32-dimensional embedding vector is retained. The objective of the proposed digital biomarker is to quantify the relative position of a gait trial within the learned embedding space with respect to the healthy, neurological, and orthopedic gait clusters. To establish reference points for each clinical category, class-specific prototype vectors are computed using the normalized embeddings of all training samples belonging to each class. The prototype of each class is defined as:

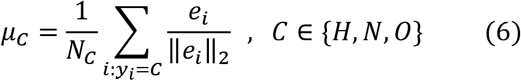

where (*N*_*C*_) denotes the number of training samples belonging to clinical classes (*H:* Healthy, *N*: Neurological, *O*: Orthopedic), (*y*_*i*_) is the class label of sample (*i*), and (*e*_*i*_) is the corresponding embedding vector. Consequently, three prototype vectors are obtained, representing the centers of the healthy (*μ*_*H*_), neurological (*μ*_*N*_), and orthopedic (*μ*_*O*_) clusters in the embedding space. For a new gait trial with embedding vector (*e*), the normalized embedding is first computed and its Euclidean distance to each class prototype is calculated as:

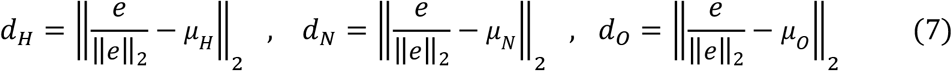

where (*d*_*H*_), (*d*_*N*_), and (*d*_*O*_) represent the distances to the healthy, neurological, and orthopedic prototypes, respectively. These distances provide an interpretable description of the similarity between a gait trial and each clinical category. The proposed embedding-distance digital biomarker is then defined as:

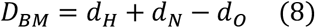

This formulation is empirically selected to maximize separation between the three clinical groups. Intuitively, the biomarker increases when the embedding moves closer to the orthopedic prototype and decreases when it moves toward the healthy or neurological prototypes. As a result, the biomarker transforms the multidimensional embedding space into a single scalar value while preserving clinically relevant information regarding gait abnormalities. The resulting digital biomarker can subsequently be used for group discrimination, statistical analysis, and reliability assessment across repeated gait trials.

## 4. Experiments and Results

The proposed network was trained using a learning rate of 1e-3, a weight decay of 1e-3, and the Adam optimizer. The number of network parameters obtained is equal to 20,532. The dataset was split into 80% for training and 20% for testing. All the experiments of this study are implemented based on PyTorch 2.11.0 library on a system with AMD Ryzen 5 5600X 3.70 GHz CPU and GeForce RTX 3060 Ti GPU.

### 4.1 Visualization of Embedding Representation

To investigate the discriminative capability of the learned representation space, the 32-dimensional embedding vectors generated by the proposed network after training are visualized using Principal Component Analysis (PCA). PCA is a linear dimensionality reduction technique that projects high-dimensional data onto a lower-dimensional space while preserving the maximum possible variance. Specifically in this technique, the covariance matrix of the embedding vectors is computed, followed by eigenvalue decomposition. The two eigenvectors associated with the largest eigenvalues are selected as the principal components, and the original embeddings are projected onto these directions. This procedure generated a two-dimensional representation that preserves the largest proportion of variance in the data and enables visual assessment of cluster formation among the clinical categories.

Fig. 3. illustrates the PCA projections of the embedding vectors obtained from both the training and test sets. As shown in visualization of training samples, the training embeddings form three well-defined clusters corresponding to the healthy, neurological, and orthopedic categories. This clustering behaviour indicates that the supervised contrastive learning successfully learned discriminative representations by minimizing intra-class variability while maximizing inter-class separation. The orthopedic and neurological groups form a particularly compact cluster, whereas the healthy group exhibit slightly larger dispersion, reflecting the greater variability of gait patterns in this group.

**Figure 3.**
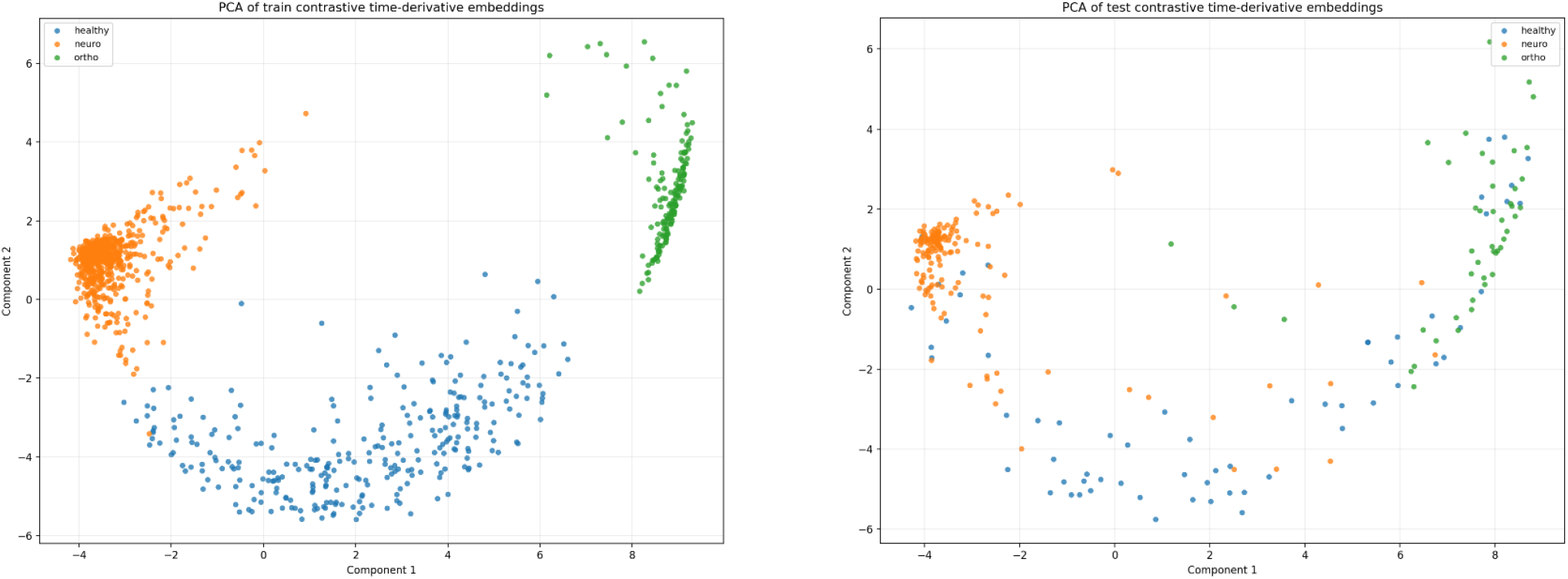
Embedding visualization after dimensionality reduction with PCA for the training (left) and test (right) samples of healthy, neurological, and orthopedic subjects

A similar clustering structure can also be observed in the unseen test subjects shown in Fig. 3. (right). Despite the increased dispersion expected from unseen participants, the three clinical categories remain acceptably separated, demonstrating the generalization capability of the proposed representation learning framework. Only a limited number of samples appear near cluster boundaries, suggesting that the learned embedding space preserved clinically relevant gait characteristics in test subjects.

The clear separation between healthy, neurological, and orthopedic gait patterns provides further justification for the prototype-based digital biomarker proposed in Section 3.4. Since samples belonging to the same clinical category naturally form compact regions within the embedding space, distances to the corresponding class prototypes can be used as meaningful measures of gait similarity. The PCA visualization therefore offered qualitative evidence that the learned embedding space is suitable for constructing a discriminative digital gait biomarker.

### 4.2 Quantitative Evaluation on Test Subjects

To quantitatively evaluate the proposed embedding-based digital biomarker, a nearest-prototype classification experiment was performed in the learned embedding space. Since class-specific centroids are obtained from the training embeddings of the healthy, neurological, and orthopedic groups, each test subject can be assigned to the class corresponding to its nearest centroid based on Euclidean distance. The predicted label is then compared with the true clinical category to assess the discriminative capability of the learned representation. This experiment is conducted on the held-out test set, which consisted only of subjects not included during training. This subject-level evaluation strategy is used to prevent information leakage across repeated trials and to assess the generalization capability of the learned gait representation. The evaluation metrics considered for each class are precision, recall, and F1-score, defined as:

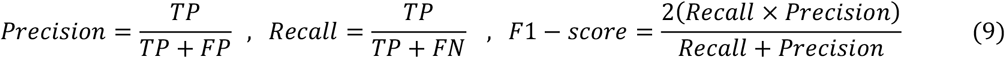

For each clinical category, these metrics are computed using a one-versus-rest strategy. True Positives (TP) denote samples correctly assigned to the corresponding class, while False Positives (FP) denote samples incorrectly assigned to that class despite belonging to another category. False Negatives (FN) denote samples belonging to the corresponding class but incorrectly assigned to a different class. True Negatives (TN) denote samples from other classes that are correctly not assigned to the class under consideration. Overall accuracy is computed as the proportion of correctly classified samples among all test subjects:

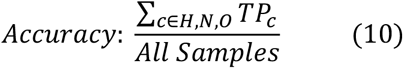

Fig. 4. presents the confusion matrix obtained on the independent test set using nearest-prototype classification in the learned embedding space. The diagonal elements represent correctly classified samples, whereas the off-diagonal elements correspond to misclassifications. The neurological category achieved the highest classification performance, with 144 out of 157 samples correctly identified. Similarly, the orthopedic category demonstrated strong discriminative performance, with 43 of 46 samples correctly classified. In contrast, the healthy class showed a lower number of correctly classified samples, with 36 out of 63 trials correctly identified. Most misclassifications of healthy samples occurred toward the neurological and orthopedic categories, with 11 and 16 samples, respectively.

**Figure 4.**
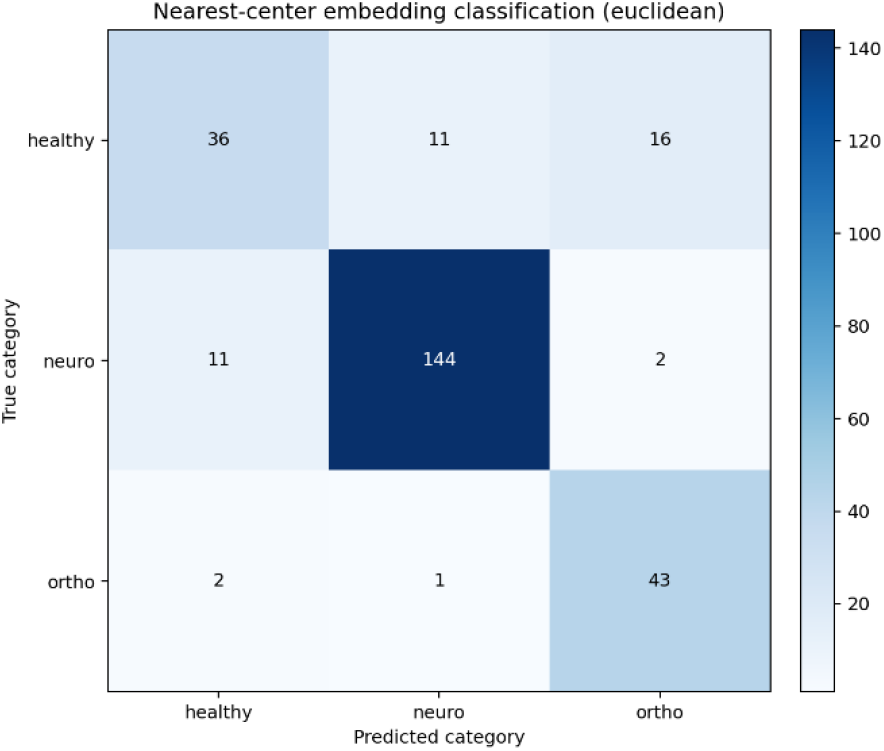
Confusion matrix for test subjects based on the Euclidian distance from centroids of healthy, neurological, and orthopedic embeddings

As shown in Table 1, the proposed digital biomarker achieved an overall accuracy of 84% on the test set. The neurological class showed the strongest performance, with precision, recall, and F1-score all equal to 0.92. This indicates that the model is highly effective in identifying neurological gait patterns and produced a low number of false positive and false negative predictions for this category. The orthopedic class also achieved high recall of 0.93, suggesting that most orthopedic samples are correctly detected, although its lower precision of 0.70 indicates some misclassification of non-orthopedic samples as orthopedic. The healthy group showed a precision of 0.73 and recall of 0.57, resulting in an F1-score of 0.64, indicating that healthy trials were more difficult to distinguish from pathological gait patterns. The high F1-score for the neurological group and the high recall for the orthopedic group suggests that the learned embedding space captures clinically meaningful differences between movement disorder categories. However, the relatively lower recall for the healthy group indicates partial overlap between healthy and pathological gait representations, which may reflect natural variability in healthy walking patterns or similarity between mild impairments and normal gait.

**Table 1.** Performance quantification results for the proposed digital biomarker.

| Evaluation Metric | Healthy | Neurological | Orthopedic |
| --- | --- | --- | --- |
| **Precision** | 0.73 | 0.92 | 0.70 |
| **Recall** | 0.57 | 0.92 | 0.93 |
| **F1-score** | 0.64 | 0.92 | 0.80 |
| **Supported Data** | 63 | 157 | 46 |
| **Accuracy** | 84% |  |  |

### 4.3 Performance Comparison Between the Proposed EDGB and Handcrafted Gait Features

To assess the effectiveness of the proposed embedding-distance digital biomarker, its discriminative capability is compared with several commonly used conventional gait measures extracted from the same dataset. Specifically, average stride time, maximum stride time, stride time variability, U-turn time, and gait cadence are selected as representative temporal gait descriptors. U-turn time represents the time required for a participant to perform a 180° turn during the walking task. Gait cadence represents the average number of steps performed per unit time and reflects the rhythmic characteristics of walking.

Fig. 5. presents the distributions of the EDGB and the handcrafted gait features for test subjects across the healthy, neurological, and orthopedic groups. Visual inspection reveals that the proposed biomarker exhibits substantially greater separation between the three clinical categories than the conventional gait measures. In particular, the neurological group forms a compact cluster with low biomarker values, whereas the orthopedic group is characterized by consistently high biomarker values. The healthy group occupies an intermediate region, resulting in clear differentiation among the three populations. In contrast, the conventional gait measures demonstrate considerable overlap between clinical categories. Among them only U-turn provides better separation, however, noticeable overlap remains between the neurological and orthopedic groups. The stronger separation achieved by the EDGB can be attributed to its representation-learning framework, which integrates information from multiple IMU locations and multiple motion characteristics, including acceleration, angular velocity, jerk, and angular acceleration. Unlike conventional gait measures that describe only a single aspect of gait performance, the proposed biomarker is derived from a learned latent representation that captures complex spatial and temporal relationships within the gait signal. Consequently, it is able to summarize multidimensional gait characteristics into a single scalar measure while preserving clinically relevant information.

**Figure 5.**
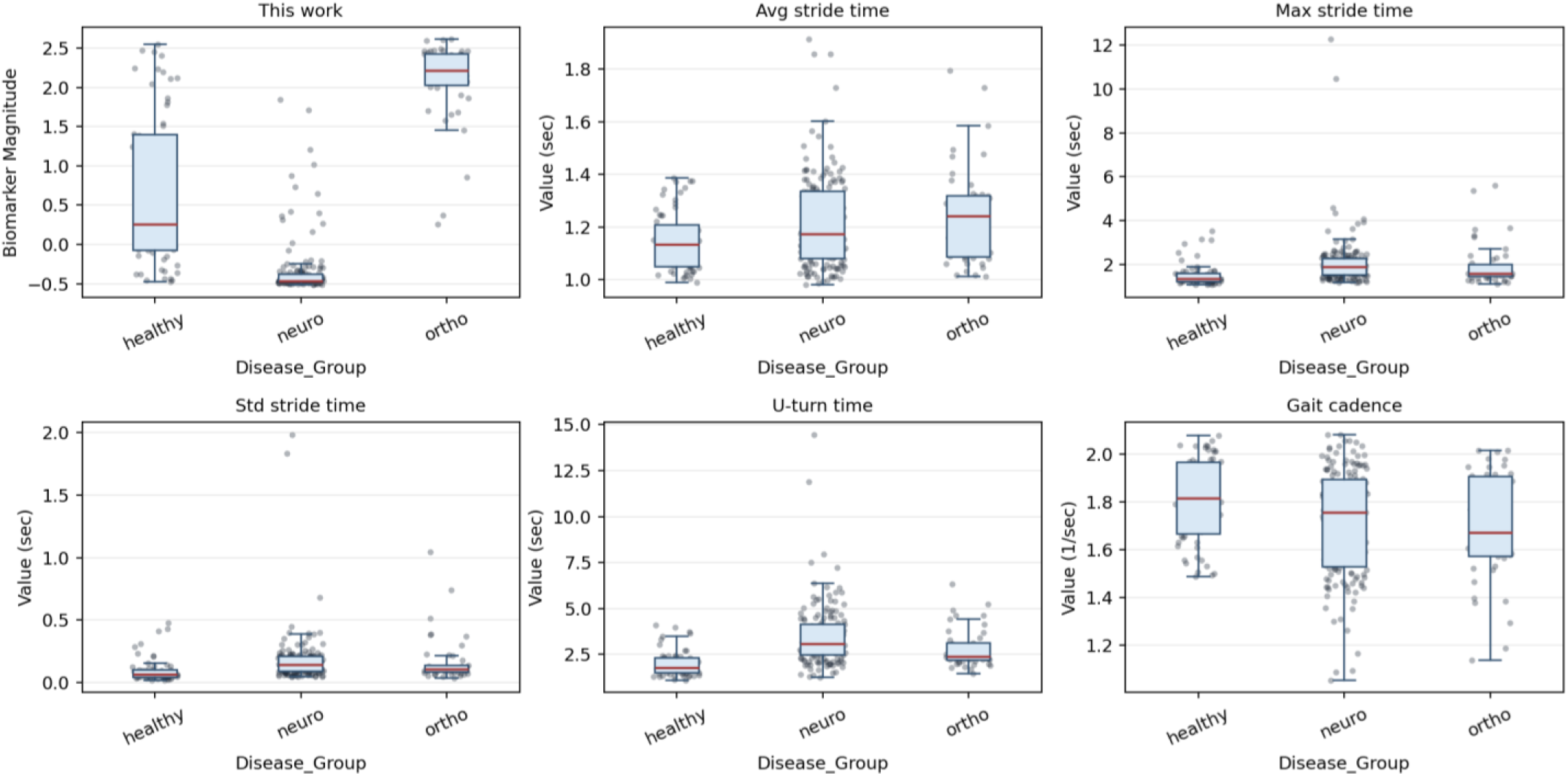
Sensitivity comparison between proposed digital biomarker and conventional gait measures (average stride time, max stride time, standard deviation of stride time, U-turn time, and gait cadence) across different disease groups for the test subjects

Another experiment used to compare the discriminative performance of the EDGB with handcrafted gait features is receiver operating characteristic (ROC) analysis. In this analysis, multiple decision thresholds are applied to each gait feature, and the corresponding true positive rate and false positive rate are computed. This makes it possible to evaluate how well each feature separates two clinical categories independently of a fixed classification threshold. Fig. 6. presents the ROC curves for the pairwise comparisons of healthy versus neurological, healthy versus orthopedic, and neurological versus orthopedic groups. The area under the ROC curve (AUC) can be used as an evaluation metric of discrimination performance, where larger values indicate better separation between clinical categories. As shown in Table 2, the EDGB consistently achieved the highest AUC values across all pairwise classification tasks. For distinguishing healthy from neurological participants, the proposed biomarker achieved an AUC of 0.9059, substantially outperforming the best handcrafted feature, U-turn time (AUC = 0.8515). Similarly, for healthy versus orthopedic discrimination, the proposed biomarker achieved an AUC of 0.8847. The largest performance difference is observed for neurological versus orthopedic discrimination where the proposed biomarker achieved an AUC of 0.9950, indicating near-perfect separation between the two pathological gait categories. In contrast, the conventional gait measures exhibited considerably lower performance, with AUC values ranging from 0.5108 to 0.6610. By averaging the AUC for all comparisons, a macro AUC of 0.9285 for EDGB is achieved which showed 17.6% improvement compared to the highest macro AUC of conventional gait measures (U-turn with AUC of 0.7648). This finding suggests that the learned representation successfully captures subtle gait characteristics specific to neurological and orthopedic disorders that are not adequately represented by conventional handcrafted descriptors.

**Figure 6.**
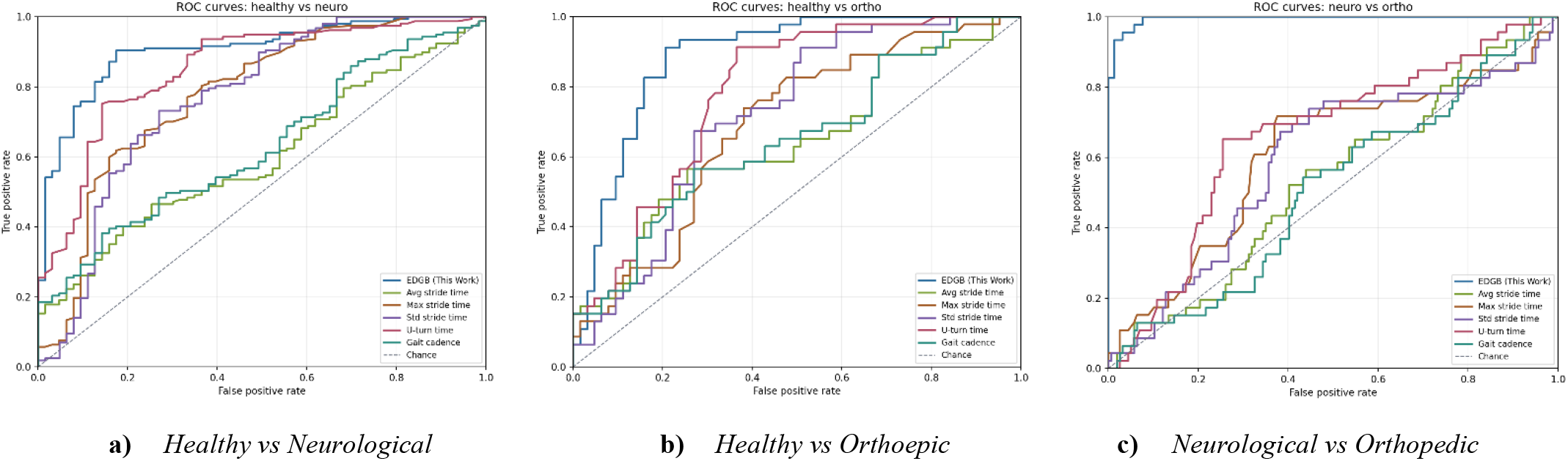
ROC curves for discriminating between multiple clinical groups

**Table 2.** AUC Results based on the gait features for comparing different clinical groups.

| Biomarker | AUC |  |  |  |
| --- | --- | --- | --- | --- |
|  | Healthy vs Neurological | Healthy vs Orthopedic | Neurological vs Orthopedic | Macro |
| Avg stride time | 0.5990 | 0.6422 | 0.5370 | 0.5927 |
| Max stride time | 0.7771 | 0.6870 | 0.6092 | 0.6911 |
| Std stride time | 0.7674 | 0.7198 | 0.5926 | 0.6933 |
| Gait Cadence | 0.6278 | 0.6479 | 0.5108 | 0.5955 |
| U-turn time | 0.8515 | 0.7819 | 0.6610 | 0.7648 |
| EDGB (this work) | 0.9059 | 0.8847 | 0.9950 | 0.9285 |

### 4.4 Ablation Study on input and sensor importance

An ablation study is conducted to investigate the contribution of individual IMU locations and input channels to the performance of the proposed EDGB biomarker. Specifically, two sets of experiments are performed. In the first experiment, the EDGB network is trained using signals from individual sensors separately and then their macro-AUC results are compared with the complete multi-sensor configuration. In the second experiment, a leave-one-channel-out analysis is performed to quantify the importance of individual input channels.

Table 3 summarizes the macro-AUC obtained when considering different sensor configurations. Among the individual sensors, the right foot (RF) sensor achieved the highest performance with a macro-AUC of 90.83%, followed closely by the left foot (LF) sensor with a macro-AUC of 90.24%. The lower back (LB) sensor achieved a macro-AUC of 88.23%, while the head-mounted (HE) sensor produced the lowest performance with a macro-AUC of 84.69%. These results indicate that foot-mounted sensors contain the most discriminative information for distinguishing healthy, neurological, and orthopedic gait patterns. To evaluate the contribution of derivative channels, the network is also trained using only raw acceleration and angular velocity signals from all four sensors. This configuration achieved a macro-AUC of 89.64%. When jerk and angular acceleration signals are additionally incorporated, the macro-AUC increased to 92.85%, representing an absolute improvement of 3.21 percentage. This result demonstrates that the derivative channels provided useful complementary information and contributed substantially to the discriminative capability of the learned representation.

**Table 3.** Macro AUC results for considering sensors separately and removing the derivative input channels.

| Sensor Data Usage | Macro-AUC |
| --- | --- |
| RF only ($a_{RF}(t) + \omega_{RF}(t) + j_{RF}(t) + \alpha_{RF}(t)$) | 90.83% |
| LF only ($a_{LF}(t) + \omega_{LF}(t) + j_{LF}(t) + \alpha_{LF}(t)$) | 90.24% |
| LB only ($a_{LB}(t) + \omega_{LB}(t) + j_{LB}(t) + \alpha_{LB}(t)$) | 88.23% |
| HE only ($a_{HE}(t) + \omega_{HE}(t) + j_{HE}(t) + \alpha_{HE}(t)$) | 84.69% |
| RF+LF+LB+HE ($a_{RF+LF+LB+HE}(t) + \omega_{RF+LF+LB+HE}(t)$) | 89.64% |
| RF+LF+LB+HE ($a_{RF+LF+LB+HE}(t) + \omega_{RF+LF+LB+HE}(t) + j_{RF+LF+LB+HE}(t) + \alpha_{RF+LF+LB+HE}(t)$) | 92.85% |

To quantify the importance of individual input channels, a leave-one-channel-out analysis is performed. In each experiment, one input channel is removed while all remaining channels are retained, and the resulting decrease in macro-AUC is recorded. The results are presented in Table 4. The largest performance degradation is observed when the right foot angular velocity channel is removed, resulting in a macro-AUC decrease of 6.18%. The left foot acceleration channel produced the second largest decrease (5.84%), followed by the right foot jerk channel (5.63%) and the right foot acceleration channel (5.51%). These findings suggest that signals acquired from the foot-mounted sensors contribute most strongly to the learned gait representation. In particular, both angular velocity and derivative-based motion descriptors from the right foot appear to play a critical role in capturing gait abnormalities.

**Table 4.** Macro AUC results after removal of some input channels to show which one of them has the most contribution.

| Input Channel Removal | Macro-AUC drop | Input Channel Importance Rank |
| --- | --- | --- |
| RF angular velocity channel ($\omega_{RF}(t)$) | -6.18% | 1<sup>st</sup> |
| LF acceleration channel ($a_{LF}(t)$) | -5.84% | 2<sup>nd</sup> |
| RF jerk channel ($j_{RF}(t)$) | -5.63% | 3<sup>rd</sup> |
| RF acceleration ($a_{RF}(t)$) | -5.51% | 4<sup>th</sup> |

### 4.5 Interpretability Analysis of EDGB

To investigate the characteristics captured by the proposed EDGB and provide more interpretability, Spearman rank correlation analysis is performed between EDGB and several conventional gait measures, including average stride time, maximum stride time, stride time variability, U-turn time, and gait cadence. The heatmap correlation coefficients are illustrated in Fig. 7. As can be seen, the proposed biomarker exhibits relatively weak correlations with all conventional gait timing related measures, with absolute correlation coefficients ranging from 0.07 to 0.26. Specifically, EDGB demonstrates weak positive correlation with average stride time (rho=0.11) and weak negative correlations with maximum stride time (rho=-0.14), stride time variability (rho=-0.15), U-turn time (rho=-0.26), and gait cadence (rho=-0.07). The strongest association was observed with U-turn time; however, the magnitude of this correlation remains low, indicating that the proposed biomarker captures information beyond conventional gait timing related characteristics. A further analysis has been performed based on some other signal-related features to demonstrate a deeper description of what EDGB is capturing. These signal-related features are average jerk, maximum jerk, std of jerk, average angular acceleration, std angular acceleration, and mean and std of acceleration and angular velocity for all of HE, LB, RF, LF located sensors. The results as shown in Fig. 8. reveal that the strength of correlation between EDGB and individual signal-related features varies substantially across sensor locations. Among all sensors, the strongest associations are observed for the foot-mounted sensors. For the RF sensor, EDGB exhibited moderate positive correlations with maximum jerk (rho=0.55), jerk variability (rho=0.46), and average jerk (rho=0.40). Similarly, for the left foot (LF) sensor, moderate correlations are observed with maximum jerk (rho=0.46), jerk variability (rho=0.38), and average jerk (rho=0.32). These findings suggest that rapid temporal changes in foot motion contribute substantially to the information encoded by the proposed biomarker. The LB sensor demonstrated comparable behavior, with the strongest correlations observed for maximum jerk (rho=0.54), acceleration variability (rho=0.47), and jerk variability (rho=0.44). In contrast, the head-mounted (HE) sensor exhibited generally weaker associations, with the largest correlation observed for maximum jerk (rho=0.49) and smaller correlations for the remaining signal-related features.

**Figure 7.**
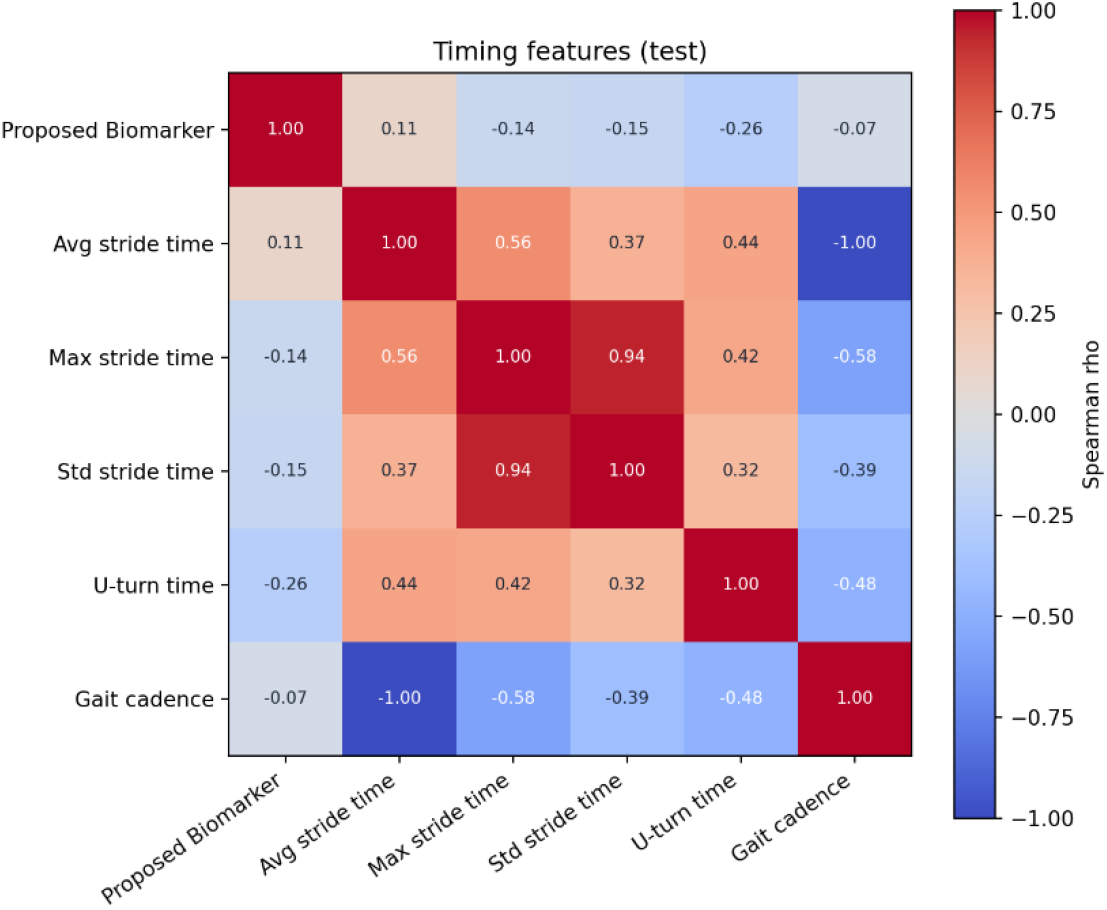
Spearman correlation heatmap between the proposed biomarker and some gait timing related conventional gait measures (average stride time, maximum stride time, std of stride time, U-turn time, and gait cadence)

**Figure 8.**
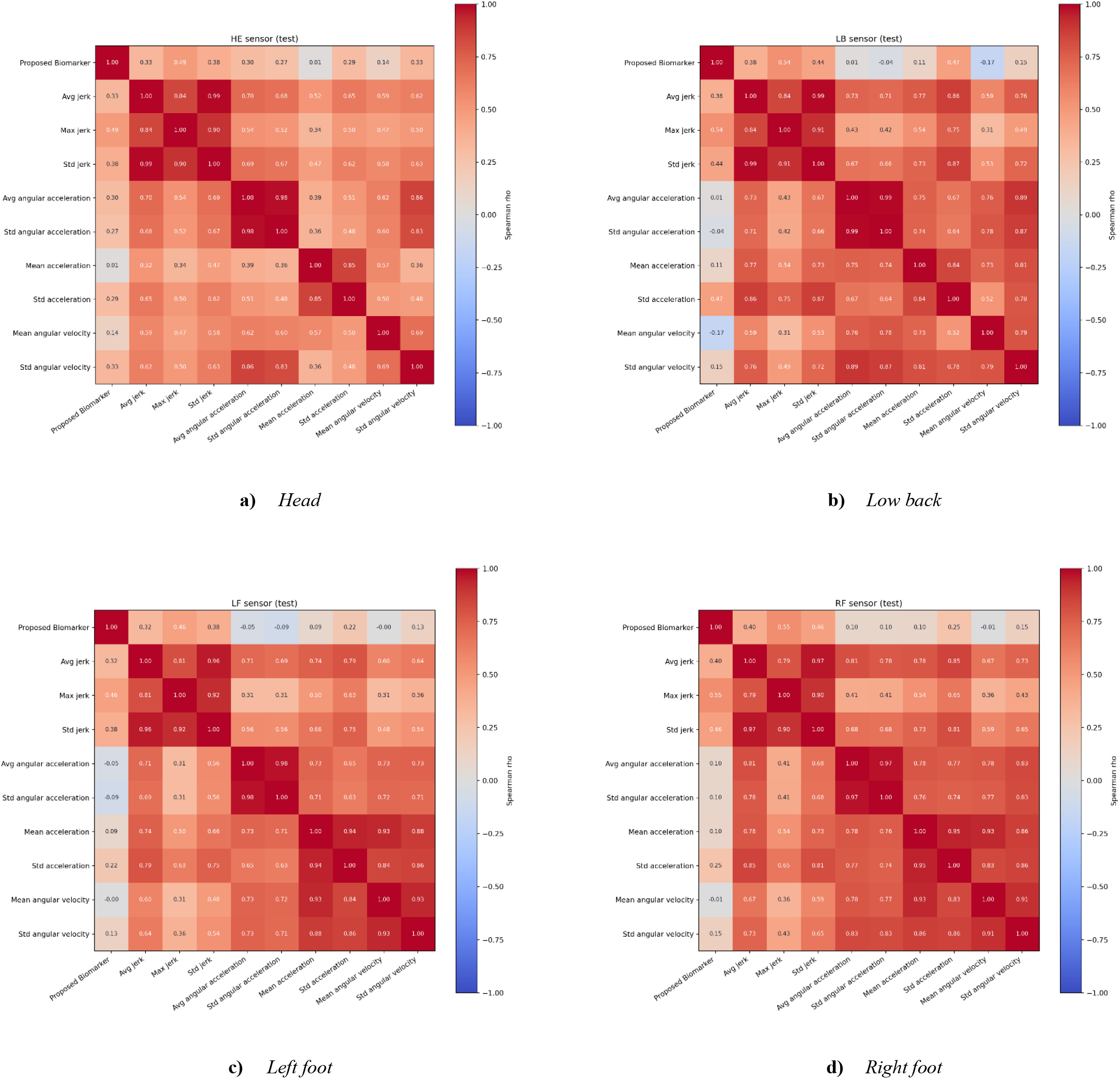
Spearman correlation heatmap between the proposed biomarker and some other signal related conventional gait measures (average jerk, maximum jerk, std of jerk, average angular acceleration, std of angular acceleration, mean and std of acceleration, mean and std of angular velocity) for head, low back, left foot, and right foot located sensors

An important observation across all four sensors is that EDGB consistently exhibits stronger associations with jerk-related measures than with conventional acceleration or angular velocity statistics. Since jerk corresponds to the temporal derivative of acceleration, it reflects rapid changes in movement and indicates that the proposed biomarker is sensitive to dynamic gait characteristics beyond simple movement magnitude.

### 4.6 Statistical Separation of Clinical Categories

In this section an Analysis of Variance (ANOVA) is performed to show statistically significance differences of the measures for the clinical categories. The result of ANOVA test is shown in Table 5, and as can be seen, the proposed EDGB demonstrated substantially stronger group separation than all conventional gait timing measures, with the largest F-statistic (322.52) and the highest effect size (η^2^ = 0.7104). This indicates that clinical category explained approximately 71% of the variance in EDGB. In comparison, U-turn time was the strongest conventional gait timing measure, with η^2^= 0.1628, while the remaining features showed relatively small effect sizes. The group means further show a clear separation for EDGB, with neurological samples having the lowest values, orthopedic samples the highest values, and healthy samples located between them.

**Table 5.** ANOVA test results for comparing the biomarker effect across healthy, neurological, and orthopedic groups.

| Biomarker | F-statistic | p-value | effect size ($\eta^2$) | Confidence Interval | Healthy mean | Neurological mean | Orthopedic mean |
| --- | --- | --- | --- | --- | --- | --- | --- |
| Avg stride time | 4.9710 | 0.0076 | 0.0364 | [0.0125-0.0864] | 1.1497 | 1.2168 | 1.2406 |
| Max stride time | 6.4289 | 0.0019 | 0.0466 | [0.0275-0.1009] | 1.5251 | 2.0947 | 1.9348 |
| Std stride time | 4.1446 | 0.0169 | 0.0306 | [0.0157-0.0856] | 0.0995 | 0.1806 | 0.1696 |
| Gait Cadence | 5.9135 | 0.0031 | 0.0430 | [0.0157-0.0993] | 1.8111 | 1.7094 | 1.6939 |
| U-turn time | 25.5632 | < 0.001 | 0.1628 | [0.1146-0.2391] | 1.9814 | 3.4773 | 2.7796 |
| EDGB (this work) | 322.5199 | < 0.001 | 0.7104 | [0.6473-0.7736] | 0.6523 | -0.3332 | 2.0994 |

### 4.7 Repeatability Assessment of EDGB

For a digital biomarker to be clinically useful, it should not only discriminate between different clinical groups but also provide consistent measurements across repeated trials from the same participant. Thanks to several trials for each subject in the dataset, a reliability analysis is performed using the Intraclass Correlation Coefficient (ICC(2,1)) to evaluate the repeatability. This metric quantifies the proportion of variability between different subjects to the total variability (include within-subject and between-subjects). Higher ICC values indicate greater consistency of measurements across repeated observations. As it has been shown in Fig. 9., ICC(2,1) value obtained for EDGB is 0.82. It indicates that 82% of the total variance in EDGB is attributable to differences between subjects. This finding suggests that the biomarker primarily reflects subject-specific gait characteristics rather than random trial-to-trial fluctuations.

**Figure 9.**
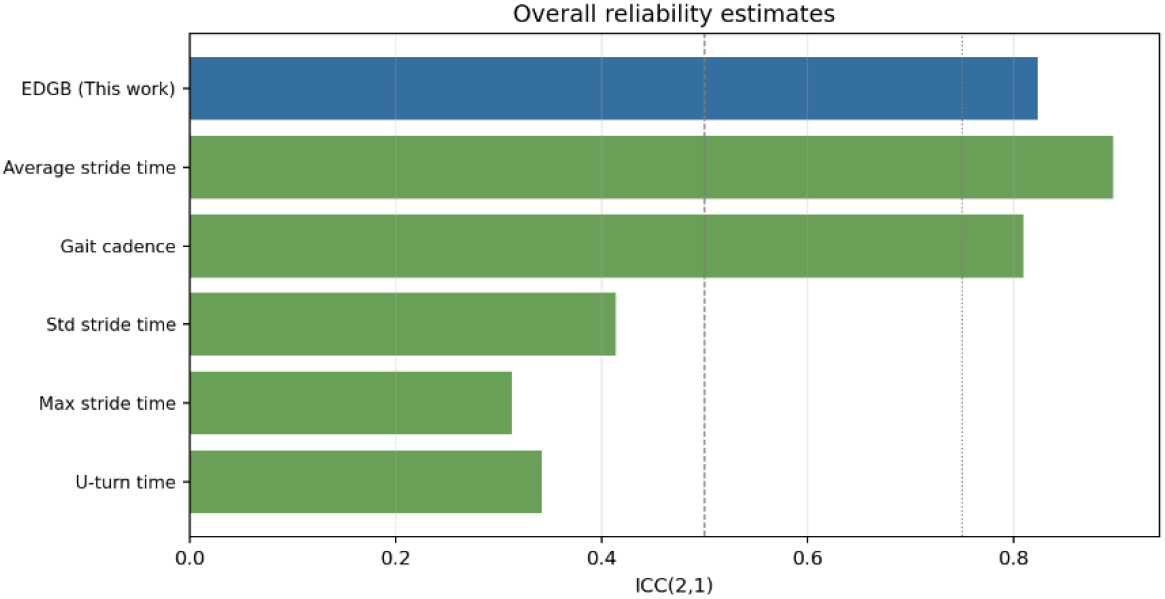
Repeatability of proposed biomarker compared to conventional gait timing measures through the ICC experiment

## 5. Discussion

The primary finding of this study is that a digital biomarker derived from a learned gait representation can substantially outperform conventional gait measures in distinguishing healthy, neurological, and orthopedic gait patterns. The proposed Embedding-Distance Gait Biomarker (EDGB) achieved a macro-AUC of 92.85%, compared with 76.48% for the best conventional gait measure (U-turn time). It suggests that the proposed biomarker captures clinically relevant gait characteristics that are not adequately represented by individual handcrafted gait measures. An important observation is that the EDGB was particularly accurate when distinguishing neurological and orthopedic gait patterns. While conventional gait measures achieved only moderate discrimination between these two pathological groups, the proposed biomarker achieved an AUC of 99.50%, indicating near-perfect separation on a held-out test subject. This result suggests that the learned representation captures subtle movement characteristics specific to different movement disorder categories. Since many conventional gait measures primarily quantify temporal aspects of walking, such as stride timing or cadence, they may fail to capture the complex spatial and dynamic information embedded within multi-sensor IMU signals. In contrast, the proposed representation-learning framework simultaneously exploits acceleration, angular velocity, jerk, and angular acceleration signals acquired from multiple body locations, enabling a richer characterization of gait abnormalities.

The spearman correlation analysis showed that the proposed biomarker appears to capture information beyond conventional gait descriptors like stride time related features and exhibited significantly stronger correlation with signal-related features such as jerk and angular acceleration. It suggests that the biomarker is not merely reproducing existing gait metrics but instead captures complementary information contained within the IMU signals. Therefore, EDGB may provide additional insight into gait impairment beyond that available from traditional gait assessment approaches.

The ablation study also revealed several practical insights. First, the highest performance among individual sensors was obtained from the foot-mounted IMUs, highlighting the importance of lower-limb kinematics in distinguishing movement disorder categories. Second, adding jerk and angular acceleration channels increased the macro-AUC from 89.64% to 92.85%. This result demonstrates that explicitly incorporating derivative signals provides valuable information that helped building compact deep learning architecture with only 20,532 parameters.

Several limitations of this study should be acknowledged. First, the proposed framework was evaluated using broad neurological and orthopedic categories and did not investigate specific disease subtypes or disease severity levels. Future work should focus on extending the learned embedding space to provide finer-grained representations capable of distinguishing between different disorders within each clinical category and capturing variations in disease severity. Furthermore, future studies should investigate the longitudinal behavior of EDGB and evaluate its sensitivity to disease progression and treatment-related changes, which are essential requirements for its application in clinical monitoring and therapeutic assessment.

Overall, the findings demonstrate that representation learning combined with supervised contrastive learning provides an effective framework for developing digital gait biomarkers. The proposed EDGB achieved superior discrimination performance, strong statistical separation, and good repeatability while maintaining a compact network architecture. These results highlight the potential of learned gait representations as objective and clinically useful biomarkers for movement disorder assessment using wearable IMU sensors.

## 6. Conclusion

This study proposed an Embedding-Distance Gait Biomarker (EDGB) based on supervised contrastive learning and multi-sensor IMU signals for the objective assessment of gait patterns in healthy, neurological, and orthopedic populations. A compact deep learning architecture consisting of one-dimensional convolutional neural network (1D-CNN) layers was developed to learn a 32-dimensional gait representation from 16 IMU-derived signals, including acceleration, angular velocity, and their corresponding derivatives (jerk and angular acceleration), acquired from the head, lower back, right foot, and left foot sensor locations. The proposed EDGB was subsequently derived from the distances between each embedding vector and class-specific prototype vectors learned during training. Experimental results demonstrated that the proposed representation-learning framework provided substantially stronger discrimination between clinical categories than conventional gait measures. interpretability analyses revealed that the biomarker captured information beyond conventional gait measures and was particularly associated with jerk-related motion characteristics, reflecting the importance of higher-order movement dynamics. This study demonstrated that supervised contrastive learning can effectively transform multi-sensor IMU recordings into a compact and informative gait representation, with potential utility for objective movement disorder assessment in both clinical and research applications.

## Data Availability

The data used in this study are publicly available through the Voisard dataset, A Dataset of Clinical Gait Signals with Wearable Sensors from Healthy, Neurological, and Orthopedic Cohorts, and can be accessed at https://doi.org/10.6084/m9.figshare.28806086. The source code developed for this study will be made publicly available through a GitHub repository upon publication of the manuscript.

## 7. Declaration of Competing Interest (Conflict of Interest)

The author declares that there are no known competing financial interests or personal relationships that could have appeared to influence the work reported in this paper.

## 8. Funding Statement

This research did not receive any specific grant from funding agencies in the public, commercial, or not-for-profit sectors.

## 9. Author Contributions

Seyed Mehdi Mohtavipour conceived the study, designed the methodology, implemented the software, conducted the experiments, analysed the results, and wrote the manuscript.

## 10. Code Availability Statement

The source code developed for this study will be made publicly available through a GitHub repository upon publication of the manuscript.

## 11. Ethics Statement

This study used the publicly available Voisard dataset, *A Dataset of Clinical Gait Signals with Wearable Sensors from Healthy, Neurological, and Orthopedic Cohorts*. The original study protocol was approved by the **Comité de Protection des Personnes Ile de France II** (Approval No. **CPP 2014-10-04 RNI**), and written informed consent was obtained from all participants prior to data collection.

## Notes

### Competing Interest Statement

The authors have declared no competing interest.

### Author Declarations

This study used the publicly available Voisard dataset, A Dataset of Clinical Gait Signals with Wearable Sensors from Healthy, Neurological, and Orthopedic Cohorts. The original study protocol was approved by the Comité de Protection des Personnes Ile de France II (Approval No. CPP 2014-10-04 RNI), and written informed consent was obtained from all participants prior to data collection.

